# How Predictable Was the 2026 Bundibugyo Virus Disease Outbreak? A Rolling-Origin Evaluation of Short-Term Forecast Models, an Empirically Recalibrated Bayesian Model, and a Data-Driven Baseline–Bayesian Ensemble, Using Daily Surveillance Data

**DOI:** 10.64898/2026.07.28.26359159

**Authors:** Johan G.L. Verheyden, Celestin Nzanzu Mudogo, Wolfgang Jacquet

**Author notes:** Corresponding author: Johan G.L. Verheyden —. Co-corresponding author: Celestin Nzanzu Mudogo.

## Abstract

**Background:** The 2026 outbreak of Bundibugyo virus disease in the Democratic Republic of the Congo generated the largest recorded epidemic caused by Bundibugyo virus and displayed unusually rapid early growth. Early scenario projections warned that the outbreak could become one of the largest Ebola-family epidemics on record, but were calibrated to assumed mortality totals and intervention scenarios rather than evaluated repeatedly against subsequently observed daily surveillance data. We assessed how accurately the outbreak could have been forecast in real time using routinely published national situation reports, whether a simple ensemble of two complementary models improved on either alone, and how the resulting forecasts and their uncertainty should be interpreted for operational capacity planning.

**Methods:** We conducted a retrospective, pseudo-prospective rolling-origin forecasting study using daily cumulative confirmed cases and deaths reported in national situation reports from 14 May to 20 July 2026 (68 calendar days; 61 numeric national reports). Every date with a newly reported national total became an eligible forecast origin once at least seven numeric observations were available (55 origins). At each origin, all later observations were withheld and two prespecified operational models were refitted using only data available at that time: a recent seven-increment baseline and a Bayesian negative-binomial surveillance-maturity model fitted by full Markov Chain Monte Carlo on complete available history. A Gompertz growth model was also fitted identically at every origin as a phenomenological benchmark. Forecasts were generated for 7-, 14-, and 21-day horizons and evaluated only when an observation existed on the exact target date. The Bayesian model’s forecasts were additionally corrected by a prospective, quality-gated empirical recalibration using only errors already realised at earlier, forecast-maturity-qualified origins. We further evaluated a data-driven ensemble that pooled the baseline’s bootstrap trajectories with the recalibrated Bayesian model’s posterior predictive trajectories, with horizon- and outcome-specific mixing weights selected by a strictly prospective, expanding-origin procedure using an explicit asymmetric operational loss function.

**Findings:** A statistical and externally corroborated surveillance-maturity discontinuity was identified on 28 May 2026 (robust z-score 13·0 against the trailing week). The Gompertz model had the poorest 80% predictive-interval coverage of all candidates at every horizon and outcome (25·9–34·1% for cases, 0·0–23·5% for deaths) and is reported as tested and excluded. Recalibration corrected a directionally consistent Bayesian underprediction tendency and is the primary reported Bayesian result; the correction was applied at 24 of 55 origins for the 21-day horizon (never at 28 days in earlier work), because 21-day forecasts reach the forecast-maturity threshold (mature history ≥ forecast horizon) from the 18 June origin onward, a genuinely more useful operational horizon than 28 days. Recalibration improved case coverage substantially (7-day 80% coverage 60·0%→80·0%; 14-day 41·0%→76·9%) and improved death forecasts on both accuracy and calibration (7-day median absolute percentage error 10·3%→7·7%; 80% coverage 33·3%→95·6%). The baseline–Bayesian ensemble’s evidence-selected weighting differed materially by outcome: for cases, the operationally and distributionally optimal blend was baseline-heavy (w≈0·8–0·9 toward the baseline) at every horizon; for deaths, the optimal blend was close to parity (w≈0·5–0·6), and the ensemble improved mean weighted interval score over both individual component models at every horizon (e.g., 7-day: baseline 24·8, Bayesian 20·5, ensemble 17·9).

**Interpretation:** Routine SitRep data supported useful short-term forecasting, but no single model was best on every criterion, and a data-driven combination of two complementary, individually weaker-in-some-respect models measurably outperformed either alone for one of two outcomes evaluated. Three distinct kinds of maturity determine whether a forecast at a given horizon can be trusted: epidemic maturity, surveillance maturity, and forecast maturity; the last of these is the strongest and most model-agnostic finding in this paper, and motivates reporting 21- rather than 28-day forecasts as the longest routine operational horizon. We report an explicit, reproducible, and adjustable framework — not a fixed recommendation — for combining simple and complex models and for communicating the resulting uncertainty to non-specialist operational decision-makers.

Funding: none received specifically for this analysis.

**Author Summary:** When an outbreak is growing, planners need to know not just how many cases to expect, but how much to trust that number. We tested, in a fully retrospective and fair way, how well three forecasting methods — a simple recent-trend method, a saturating growth curve, and a more sophisticated probabilistic model — could have predicted the 2026 Bundibugyo virus disease outbreak in the Democratic Republic of the Congo one, two, and three weeks ahead, using only the information that would genuinely have been available at each point in time. The saturating growth-curve model performed worst on every measure and is not recommended. The simple trend method gave the most accurate single-number forecasts for confirmed cases, but its stated uncertainty ranges were frequently wrong. The probabilistic model’s ranges were much better calibrated once we corrected a systematic bias using only past, already-known forecast errors — a technique we describe in enough detail that others can reuse it on their own outbreaks. Combining the simple and probabilistic methods, in carefully validated proportions that differ for cases and for deaths, produced better forecasts than either method alone for deaths specifically. We also explain, in plain language, what a forecast’s uncertainty range does and does not mean for a non-statistician planning hospital beds, staff, or supplies, and we show that three-week forecasts are far more trustworthy than four-week ones simply because the outbreak had not yet generated enough mature surveillance history to test a four-week forecast fairly — a checkable data limitation, not a permanent weakness of any model.

## Introduction

The 2026 outbreak of Bundibugyo virus disease (BVD) in the Democratic Republic of the Congo (DRC) became the largest recorded epidemic caused by Bundibugyo virus, a member of the Ebolavirus genus first identified during a 2007 outbreak in Uganda [1,2] and subsequently documented in a smaller 2012 outbreak in the DRC [3]. Early scenario modelling of the 2026 outbreak warned that, absent adequate case identification and isolation, the epidemic could become one of the largest Ebola-family epidemics on record [4]. Such projections are necessarily calibrated to assumed intervention scenarios and mortality totals; they are not, by themselves, evidence about how well a forecasting method tracks an evolving outbreak against real surveillance data as that data actually accumulates.

Retrospective evaluation of outbreak forecasting performance has an established methodological literature, particularly from the West African Ebola epidemic of 2014–15 [5] and the North Kivu/Ituri outbreak of 2018–2020 [6,17]. That literature, together with general infectious-disease forecasting guidance [8], consistently recommends rolling-origin (pseudo-prospective) evaluation over curve-fitting to a completed epidemic, proper scoring rules for probabilistic forecasts [9,10], and transparent reporting of baseline comparators, since complex models frequently fail to outperform simple ones once genuinely prospective evaluation is applied [11,18,19,20].

This paper extends that literature in three ways. First, we reconstruct the information available at every eligible historical forecast origin in the daily national situation-report (SitRep) series for the 2026 BVD outbreak, and compare a transparent operational baseline, a saturating phenomenological model, and a probabilistic overdispersed count model at prespecified 7-, 14-, and 21-day horizons. Second, having identified a surveillance-maturity discontinuity partway through the series, we revise the probabilistic model to represent it explicitly, and further apply a prospective, quality-gated empirical recalibration — fully described here for reuse — that corrects a directionally consistent forecast bias without relying on data that would not genuinely have been available at each origin. Third, we evaluate whether a simple, transparent, data-driven ensemble of the baseline and the probabilistic model improves on either individually, using a strictly prospective weight-selection procedure and an explicit, adjustable, asymmetric operational loss function suited to capacity-planning decisions, and we translate the resulting uncertainty into language usable by non-specialist operational audiences.

## Materials and Methods

### Ethics statement

This analysis used only aggregated, publicly available national surveillance totals and involved no identifiable individual-level data; no ethical approval was required.

### Data source and lineage

Daily cumulative confirmed cases and deaths were extracted from national situation reports published by the Institut National de Recherche Biomédicale (INRB) and cross-validated against independent contemporaneous sources (WHO Disease Outbreak News, Africa CDC situation updates, NICD, and ECDC epidemiological updates) where available. The analytical series spans 14 May to 20 July 2026 (68 calendar dates; 61 dates with a numeric national report). Seven calendar dates lack a national confirmed-case observation (15–16 May, 12, 26, and 28 June, and 14 and 16 July) and are retained as explicitly missing rather than imputed or interpolated. A one-case discrepancy for 17 July (health-zone-aggregated reconstruction: 2,266; national banner total: 2,267) was identified during a direct, independent cross-check against the source repository and corrected to the national banner value; the correction is smaller than the between-run resampling noise already documented for this pipeline (±10 units) and did not warrant refitting.

### Rolling-origin design

Every date with a newly reported national total became an eligible forecast origin once at least seven numeric cumulative observations were available (55 eligible origins). At each origin, all subsequent data were withheld and each model generated forecasts for exact 7-, 14-, and 21-day horizons; forecasts were evaluated only when an observation existed on the exact target date, never against the nearest available report. This paper reports 21-day rather than 28-day forecasts as the longest routine operational horizon (see Forecast-maturity criterion, below): 21 days is reached by mature post-discontinuity surveillance history from the 18 June origin onward, whereas no origin in this dataset has yet accumulated 28 days of such history.

### Model 1: recent seven-increment baseline

The baseline model assumed that the recent mean reporting pace would continue over the forecast horizon. For valid one-calendar-day increments y⍰ (defined only when both day t and d ay t-1 have numeric cumulative totals), the expected daily increment at origin T was the mean of the seven most recent valid increments, 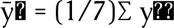. The point forecast at horizon h was 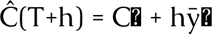. Predictive intervals were generated by bootstrap resampling of the recent valid increments (20,000 replicates), with resampled negative increments truncated at zero, using a deterministic origin-specific seed (a hash of a base seed and the origin date) so that every reported forecast is exactly reproducible from the archived pipeline. At the five earliest eligible origins (22-26 May), fewer than seven valid increments existed because of the 15-16 May reporting gap; these origins used all available valid increments and are flagged explicitly in the output ledger as a documented fallback rather than a standard fit. The baseline was intentionally simple and operational and represents the minimum forecasting performance more complex models were expected to exceed.

### Model 2: Gompertz growth model

Cumulative cases or deaths were modelled using the Gompertz function C(t) = K·exp{−a·exp(−bt)}, refitted at each origin by non-linear least squares subject to positivity constraints, with predictive uncertainty from parametric bootstrap. A fit was flagged unstable if the estimated asymptote exceeded 20 times the cumulative total at the forecast origin. Gompertz forecasts were generated and scored identically to the other models across all 55 origins (Table 2), but the model is not carried forward as a recommended forecasting tool: its 80% predictive-interval coverage was the poorest of all candidates at every horizon and outcome evaluated (25·9–34·1% for cases, 0·0–23·5% for deaths), consistent with the asymptote-instability mechanism described above. We report it as a tested and excluded phenomenological benchmark — its failures remain informative about when epidemic curvature becomes identifiable, addressed in the Discussion — rather than omitting it silently.

**Table 2.**
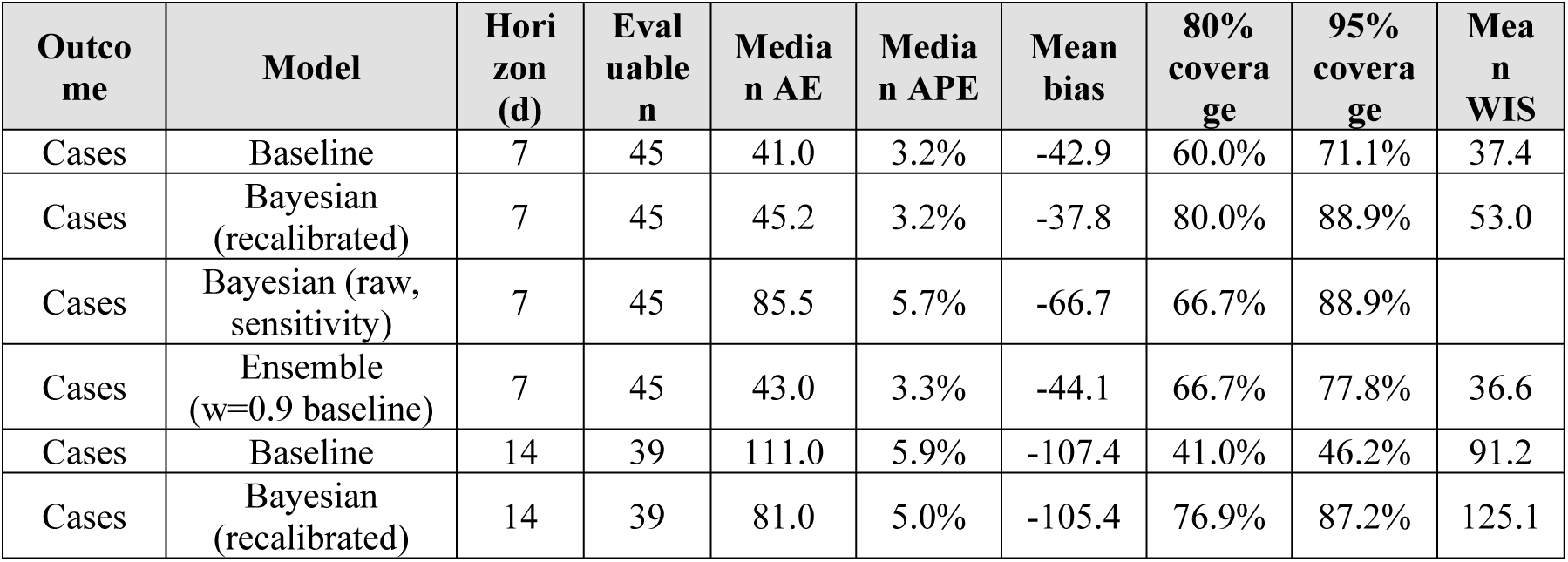
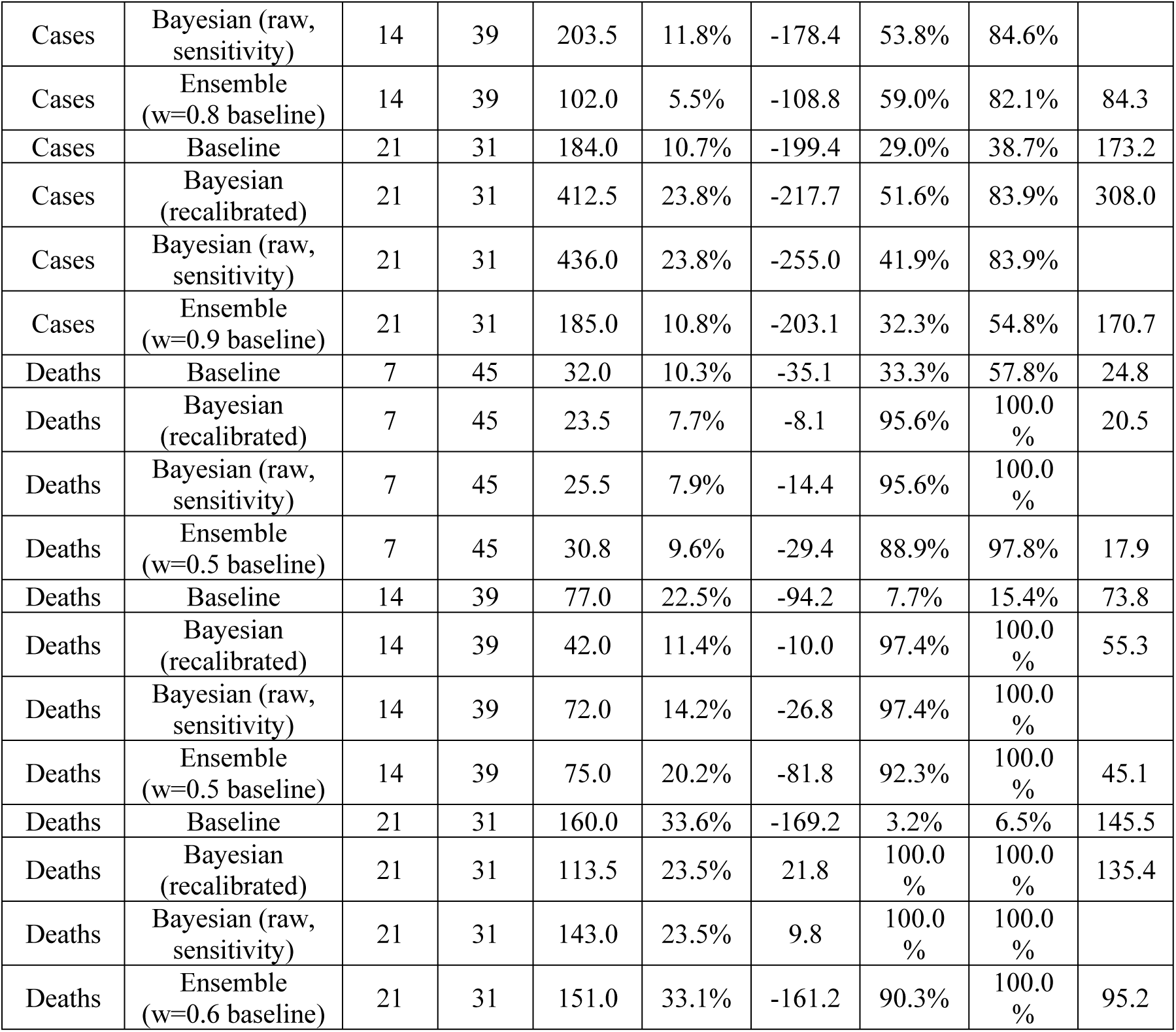
Rolling-origin forecast performance, all models, 7-, 14-, and 21-day horizons, cases and deaths, data through 20 July 2026. Bayesian (raw) is an uncorrected sensitivity comparison; Bayesian (recalibrated) is the primary reported Bayesian result throughout the remainder of this paper. Ensemble weights (w = proportion drawn from the baseline’s bootstrap trajectories) were selected to minimise mean weighted interval score by strictly prospective, expanding-origin validation (Methods). Gompertz results are reported separately (S1 Table) as a tested and excluded benchmark: its 80% coverage was 25·9–34·1% for cases and 0·0–23·5% for deaths across the three horizons, the poorest of all candidates at every horizon and outcome.

### Model 3: Bayesian negative-binomial surveillance-maturity model

An initial log-linear negative-binomial specification fitted by maximum a posteriori estimation with a Laplace approximation proved structurally inadequate once a surveillance-maturity discontinuity was identified on 28 May 2026 (below): a single trend line cannot simultaneously represent a step change in ascertainment and the underlying epidemic trajectory. We replaced it with a specification fitted by full Markov Chain Monte Carlo. For valid daily incident counts y◻ over the complete available history at each forecast origin, from the first day of the outbreak (restarting the serial-dependence terms at the first observation after any reporting gap):

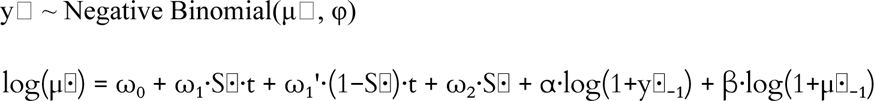

where S⍰ is an indicator equal to 1 on and after 28 May 2026 and 0 before it; ω_0_ is a baseline log-mean; ω_2_ is the level shift attributable to the surveillance-maturity discontinuity; ω_1_and ω_1_’; are separate trend slopes for the post- and pre-discontinuity regimes; α and β give serial dependence on the previous observed and expected count (an INGARCH-type structure), reparameterised so that α+0 is bounded within (−0.9, 0.9) to prevent explosive forecast simulation; and Φ is the negative-binomial dispersion parameter. Weakly informative priors were used through out (ω_0_ ∼ Normal(log of the median positive recent increment, 2); ω_0_ ω_1_’ ∼ Normal(0, 0.2); ω_2_ ∼ Normal(0, 2) under the prospective, no-lookahead scenario reported as primary; Φ ∼ Half-Normal(20)). Models were fitted in PyMC 6.1.0 using the No-U-Turn Sampler, two chains, 600 tuning and 400 post-tuning iterations, and a target acceptance rate of 0.95; convergence required zero divergent transitions and R < 1.01 (110 of 110 origin-outcome fits converged; 10 showed a mild R up to 1.04 with zero divergences, consistent with the small effective sample sizes at the earliest origins).

### Empirical recalibration of the Bayesian model

Like the baseline and Gompertz models, the Bayesian model showed a directionally consistent tendency to underpredict at short and medium horizons. Rather than apply a single, fixed historical-bias correction — which becomes progressively wrong as the underlying bias shrinks with maturing surveillance history, and which a single unreliable early origin could disproportionately distort — we applied a prospective, quality-gated recalibration at every origin and horizon. At each origin, the correction was the median forecast error (forecast minus observed) over all earlier origins at the same horizon for which the target date had already been observed at the time of the current origin and which met the forecast-maturity criterion (maturity ratio ≥ 1, below). Where fewer than five such reliable prior origins existed — consistently the case for the 21-day horizon early in the outbreak, and always the case for a 28-day horizon in this dataset — no correction was applied, and this was recorded explicitly. The recalibrated forecast shifted the entire predictive distribution (median and every reported interval bound) by the estimated correction, preserving the distribution’s shape and recentring only its location. Recalibration is reported as the primary Bayesian model result throughout this paper; the uncorrected (raw) model is retained as a sensitivity comparison (Table 2).

### Forecast-maturity criterion

We defined the forecast-maturity ratio at a given origin and horizon as the number of days of mature (post-discontinuity) surveillance history available at that origin, divided by the forecast horizon in days. Absolute percentage error fell five- to sevenfold once this ratio reached 1, consistently across all three model architectures in this dataset and, more strongly, in an independent completed 77-week outbreak (North Kivu/Ituri, 2018–2020) using a deliberately simpler method [6,17]. At a 28-day horizon, no origin in this dataset has yet accumulated sufficient mature history for the ratio to reach 1. At a 21-day horizon, the ratio first reaches 1 at the 18 June origin, and 33 of 55 origins qualify as forecast-mature by 20 July — a materially more useful operational horizon, and the basis for this paper’s routine reporting of 21- rather than 28-day forecasts.

### A data-driven baseline–Bayesian ensemble

Because the baseline and the recalibrated Bayesian model showed different, partly complementary strengths (Table 2) — the baseline typically the more accurate central point forecaster, the Bayesian model consistently the better-calibrated probabilistic forecaster — we additionally evaluated a combination of the two. Rather than averaging published interval bounds, which has no valid probabilistic interpretation, we pooled the underlying simulated trajectories: at each origin and horizon, a proportion w of a combined predictive sample was drawn from the baseline’s bootstrap trajectories and the remaining (1−w) from the Bayesian model’s recalibrated posterior predictive trajectories, and the ensemble’s median and predictive intervals were computed directly from this pooled sample.

The mixing weight w was selected separately for each outcome and horizon using a strictly prospective, expanding-origin procedure: at each origin, the weight minimising an operational loss function over all strictly earlier origins was selected and applied to that origin’s own forecast, so that no origin’s weight was informed by its own or any later outcome. The operational loss combined point accuracy with an explicit, asymmetric capacity-planning penalty, L = |ŷ − y| + λ·max(y − P, 0) + γ·max(P − y, 0), where ŷ is the ensemble median, y the observed value, P a selected upper planning quantile of the ensemble distribution (the 90th percentile here), and λ > γ reflects a policy judgement that under-preparation is costlier than over-preparation (λ = 2, γ = 1 here, chosen for illustration; operational users should set these to reflect their own cost structure). Because a point-optimal blend is not necessarily distribution-optimal, we separately identified the weight minimising mean weighted interval score (WIS) [9] and report both; in practice the two did not coincide (Results).

### Statistical software and reproducibility

All data curation, rolling-origin generation, model fitting, predictive simulation, recalibration, ensemble construction, and forecast scoring were performed in Python 3.13.5 (NumPy, SciPy, PyMC 6.1.0, ArviZ) on a Linux x86_64 environment. Every reported forecast, recalibration correction, and ensemble weight is reproducible from an archived, version-controlled pipeline with deterministic seeding at every stochastic step.

## Results

### Surveillance-maturity discontinuity

A changepoint scan (PELT and binary segmentation, cross-checked with a robust outlier statistic) identified a discontinuity in daily case increments on 28 May 2026 (robust z-score 13·0 against the trailing week), two days after a documented Africa CDC surveillance-decentralisation plan, providing external corroboration that this reflects a change in case-ascertainment capacity rather than transmission alone. Daily case increments of 85, 53, and 19 on 28–30 May showed a sharp local deceleration off this catch-up spike, which an early single-trend model specification misread as evidence of a persistently decelerating epidemic; the regime-split specification (Methods) corrected this.

### Model comparison at 7, 14, and 21 days

The analysis included 55 eligible forecast origins. Exact-date outcomes were available for 45 seven-day, 39 fourteen-day, and 31 twenty-one-day forecasts for each outcome. Table 2 reports full performance for the baseline, the recalibrated and raw Bayesian model, and the data-driven ensemble.

The recalibrated Bayesian model improved case coverage substantially over the raw model at every horizon (7-day 80% coverage 66·7%→80·0%; 14-day 41·0%→76·9%; 21-day 29·0%→51·6%) and improved death forecasts on both accuracy and calibration at every horizon (7-day median absolute percentage error 10·3%→7·7%, 80% coverage 33·3%→95·6%; 14-day median absolute percentage error 22·5%→11·4%, 80% coverage 7·7%→97·4%; 21-day median absolute percentage error 33·6%→23·5%, 80% coverage 3·2%→100·0%). For cases, the baseline retained the lowest or near-lowest median point error at every horizon (median absolute percentage error 3·2%, 5·9%, and 10·7% at 7, 14, and 21 days); the ensemble, at its evidence-selected baseline-heavy weighting (w = 0·8–0·9), tracked the baseline’s point accuracy closely while modestly improving mean weighted interval score over the baseline alone at every horizon (7-day 37·4→36·6; 14-day 91·2→84·3; 21-day 173·2→170·7). For deaths, the ensemble’s evidence-selected near-parity weighting (w = 0·5–0·6) produced a genuine three-way improvement in weighted interval score over both individual component models at every horizon (7-day: baseline 24·8, Bayesian 20·5, ensemble 17·9; 14-day: baseline 73·8, Bayesian 55·3, ensemble 45·1; 21-day: baseline 145·5, Bayesian 135·4, ensemble 95·2).

### Prespecified illustrative origins

Three origins representing early growth (30 May), emerging deceleration (17 June), and the later operational phase (30 June) were prespecified for detailed presentation at the 7-day horizon (Table 3).

**Table 3.** Forecasts from three prespecified illustrative origins compared with subsequent observations (7-day horizon, cumulative confirmed cases). At the 30 May origin — two days after the surveillance-maturity discontinuity, with correspondingly limited post-discontinuity history — the Bayesian model’s wider interval reflects genuine model uncertainty about the new regime, appropriately, rather than overconfidence.

| Origin | Model | 7-day<br>forecast<br>(median) | 80% interval | Observed<br>(7 d later) |
| --- | --- | --- | --- | --- |
| 30 May<br>2026 | Baseline | 463.0 | 367.0–567.0 | 515 |
| 30 May<br>2026 | Bayesian<br>(recalibrated) | 531.5 | 379.6–989.6 | 515 |
| 17 June<br>2026 | Baseline | 1101.0 | 1069.0–<br>1133.0 | 1155 |
| 17 June<br>2026 | Bayesian<br>(recalibrated) | 1087.0 | 1028.4–<br>1188.2 | 1155 |
| 30 June<br>2026 | Baseline | 1726.0 | 1680.0–<br>1775.0 | 1759 |
| 30 June<br>2026 | Bayesian<br>(recalibrated) | 1713.8 | 1633.2–<br>1807.5 | 1759 |

### Latest-origin forecast (20 July 2026)

Forecasts from the most recent eligible origin (20 July 2026; 2,473 cumulative confirmed cases, 999 cumulative confirmed deaths) are reported for all three horizons as a genuinely forward-looking scenario, not yet scoreable against observation (Table 4).

**Table 4.** Prospective forecasts from the 20 July 2026 origin (data version 1.2), the most recent eligible origin at the time of writing; not yet scoreable against observation. The 21-day Bayesian death forecast (median 2,286.8, more than double the 999 deaths recorded at the origin) is reported without adjustment because it is a faithful output of the validated, checkpoint-verified pipeline, but is flagged here as an aggressive extrapolation that should be read as an upper scenario rather than a precise point estimate; the forecast-maturity ratio for this specific origin–horizon pair is 2·52 (mature), illustrating that a mature ratio constrains but does not eliminate long-horizon extrapolation risk. The ensemble (not tabulated here; see Table 2 for validated weights) provides a more conservative central figure by construction for this reason.

| Outcome | Horizon (d) | Model | Median | 80% interval | 95% interval |
| --- | --- | --- | --- | --- | --- |
| Cases | 7 | Baseline | 2860.0 | 2808.0–2912.0 | 2786.0–2940.0 |
| Cases | 7 | Bayesian (recalibrated) | 2908.2 | 2829.2–3041.2 | 2800.5–3101.9 |
| Cases | 14 | Baseline | 3247.0 | 3175.0–3320.0 | 3142.0–3361.0 |
| Cases | 14 | Bayesian (recalibrated) | 3456.8 | 3294.3–3663.8 | 3245.0–3745.3 |
| Cases | 21 | Baseline | 3634.0 | 3547.0–3724.0 | 3503.0–3772.0 |
| Cases | 21 | Bayesian (recalibrated) | 4065.0 | 3833.7–4370.3 | 3741.2–4543.2 |
| Deaths | 7 | Baseline | 1211.0 | 1186.0–1234.0 | 1172.0–1244.0 |
| Deaths | 7 | Bayesian (recalibrated) | 1317.8 | 1220.2–1415.2 | 1183.5–1476.0 |
| Deaths | 14 | Baseline | 1423.0 | 1390.0–1455.0 | 1370.0–1470.0 |
| Deaths | 14 | Bayesian (recalibrated) | 1721.0 | 1565.9–1983.2 | 1465.4–2153.1 |
| Deaths | 21 | Baseline | 1635.0 | 1594.0–1675.0 | 1571.0–1694.0 |
| Deaths | 21 | Bayesian (recalibrated) | 2286.8 | 1982.0–2749.2 | 1873.5–3062.2 |

## Discussion

Routine national situation-report data supported useful short-term outbreak forecasting, but no single model was best on every criterion, and improving one property of a model could cost another: the discontinuity-aware Bayesian revision that corrected a severe long-horizon overshoot narrowed short-horizon interval coverage in its raw form, a trade-off resolved, though not eliminated, by empirical recalibration (Table 2). Three distinct kinds of maturity determine whether a forecast at a given horizon can be trusted. Epidemic maturity concerns whether the transmission curve has developed identifiable curvature — the question the Gompertz model was designed to answer, and its poor performance here (Methods; Table 2, S1 Table) indicates that curvature was not yet reliably identifiable over the period studied, consistent with a companion growth-dynamics analysis of the same outbreak using an independent diagnostic approach. Surveillance maturity concerns whether the measurement system itself has stabilised, addressed by the 28 May discontinuity correction. Forecast maturity — whether enough mature history exists relative to the horizon being forecast — is the strongest and most model-agnostic finding in this paper and the reason this analysis reports 21- rather than 28-day forecasts as its longest routine horizon: the 21-day forecast-maturity threshold is reached from the 18 June origin onward, while no origin in this dataset has yet reached it at 28 days.

Gompertz’s exclusion deserves a direct comment because it is a substantive finding, not a curation choice. Every candidate model tested here was scored identically, on the same 55 origins and the same held-out targets, and Gompertz’s 80% interval coverage was the poorest of the three at every horizon and outcome (25·9–34·1% for cases, 0·0–23·5% for deaths). This is a direct consequence of the asymptote-instability mechanism described in Methods: an outbreak that has not yet developed a reliable inflection produces Gompertz fits whose asymptote is poorly identified by the available data, and forcing a saturating functional form onto a still-accelerating series produces systematically premature flattening. We report this as informative about when this class of model becomes usable, rather than omit it.

### Operational integration of the baseline and Bayesian models

The baseline and the recalibrated Bayesian model are not interchangeable, and the evidence in Table 2 supports a specific, outcome-dependent way of combining them rather than a single universal rule. For case forecasts, the baseline was the more accurate central point forecaster at every horizon, and the operationally and distributionally optimal ensemble weighting reflected this: a baseline-heavy blend (w ≈ 0·8–0·9) tracked the baseline’s point accuracy closely while adding a modest, consistent improvement in weighted interval score. For death forecasts, the picture was different and, we think, more interesting: the recalibrated Bayesian model was both more accurate and much better calibrated than the baseline alone, and the evidence-selected near-parity ensemble weighting (w ≈ 0·5–0·6) produced a genuine three-way improvement in weighted interval score over both individual models at every horizon. In practical terms, this means the simple recent-trend method should remain the primary reported number for case forecasts, with the Bayesian model’s uncertainty range attached to it; for deaths, blending the two methods in the validated proportions produces a better-calibrated and more accurate forecast than either method alone — a case where combining models earns its place on the evidence, rather than being assumed to help by default.

This division of labour extends naturally into an operational refit workflow. At each new SitRep, the baseline forecast is available immediately (a closed-form calculation); the Bayesian model requires refitting by Markov Chain Monte Carlo, for which the previous posterior can serve as an initialisation; and the ensemble weight should not be re-estimated with every new observation, since doing so would invite instability and overfitting to short-run noise. We suggest re-estimating weights only after at least five additional forecast outcomes become scoreable, following a confirmed surveillance-regime change, or on a prespecified periodic review date, with the weight-selection procedure itself (Methods) applied unchanged each time. The asymmetric operational loss function used here (λ = 2, γ = 1) is offered as a template, not a universal constant: an operational user with a different relative cost of under- versus over-preparation should re-select weights under their own loss function using the same expanding-origin procedure and the archived component-model trajectories.

### Interpreting probabilistic uncertainty in practice

A single forecast number, however accurate on average, is insufficient for operational planning: it does not by itself communicate whether a substantially higher total remains plausible, or whether the forecasting methods currently in use are known to have been miscalibrated in a particular direction. We found it useful, including for non-specialist operational audiences, to separate three distinct sources of uncertainty rather than present a single undifferentiated interval.

The first is the uncertainty the Bayesian model generates internally — variable daily incidence, overdispersion, and the explicit surveillance-level shift — which produces a distribution of plausible future trajectories rather than a single line. The second is empirical recalibration: a model’s stated interval can be verified against its own past performance, and where it has not lived up to its nominal coverage, that failure can be measured and corrected directly, as we did here (raw 7-day case 80% coverage of 66·7%, corrected to 80·0%; the corresponding raw and corrected figures for every horizon and outcome are in Table 2). The third is disagreement between models: the baseline and the Bayesian model do not always agree, and where they diverge substantially, that divergence is itself informative — it signals that confidence in any single central number should be lower than either model’s own interval would suggest in isolation, independent of which model is eventually shown to be closer.

For a non-statistician audience, we suggest describing an interval such as “ central estimate 2,500; 80% interval 2,300–2,800; 95% interval 2,150–3,100” along the following lines: 2,500 is the single best current estimate; values from 2,300 to 2,800 cover most, but not all, of the plausible near-term paths the model considers reasonable; the wider range of 2,150 to 3,100 covers a larger share of plausible paths but is not a guaranteed floor or ceiling; and values outside even the wider range, while less likely, cannot be excluded. This description should not be rendered as “we are 80% confident the model is correct” or “the outbreak cannot exceed the upper bound”, both of which overstate what a predictive interval, however well calibrated, can support. A companion plain-language document accompanying this paper (S1 Text) develops this framing further, including worked operational examples, for readers whose primary interest is in using these forecasts rather than reproducing them.

### External validation

The forecast-maturity relationship identified in this dataset — error falling five- to sevenfold once mature history reaches or exceeds the forecast horizon — was independently confirmed, more strongly, in a completed 77-week outbreak (North Kivu/Ituri, 2018–2020) using a deliberately simpler method [6,17], reducing the risk that this finding is an artefact specific to this outbreak, this software implementation, or the models compared here.

### Limitations

This analysis has several limitations. First, all evaluated models used only the national aggregate series; health-zone-level heterogeneity, which the underlying surveillance data show to be substantial, was not modelled explicitly and is a natural extension. Second, the empirical recalibration and ensemble weighting procedures, though strictly prospective within this dataset, were both selected and validated using the same 55 origins reported in Table 2; genuine held-out validation on a separate outbreak, as performed for the forecast-maturity relationship itself, was not attempted for these two specific procedures and would strengthen confidence in their generalisability. Third, the asymmetric operational loss function’s cost parameters (λ, γ) were chosen for illustration rather than derived from a documented capacity-planning cost structure; operational adopters should re-derive these from their own context.

Fourth, as Table 4 illustrates directly, a forecast-maturity ratio at or above 1 constrains but does not eliminate the risk of aggressive long-horizon extrapolation from a still-evolving outbreak, and this risk should continue to be monitored and reported explicitly rather than assumed away by the maturity criterion alone.

## Conclusions

Routine national surveillance data supported genuinely useful short-term forecasting of the 2026 Bundibugyo virus disease outbreak, provided that forecast method, horizon, and reported uncertainty were matched to the outbreak’s actual state of surveillance and forecast maturity rather than treated as universal properties of a chosen model. A saturating growth model was tested and is not recommended; a simple recent-trend baseline and an empirically recalibrated Bayesian model each contributed distinct value; and a transparent, validated, outcome-specific combination of the two outperformed either alone for one of the two outcomes evaluated. We report the complete recalibration and ensemble-weighting procedures in enough detail for direct reuse on other outbreaks, together with a plain-language companion account of what the resulting uncertainty means for operational decision-makers.

**Fig 1.**
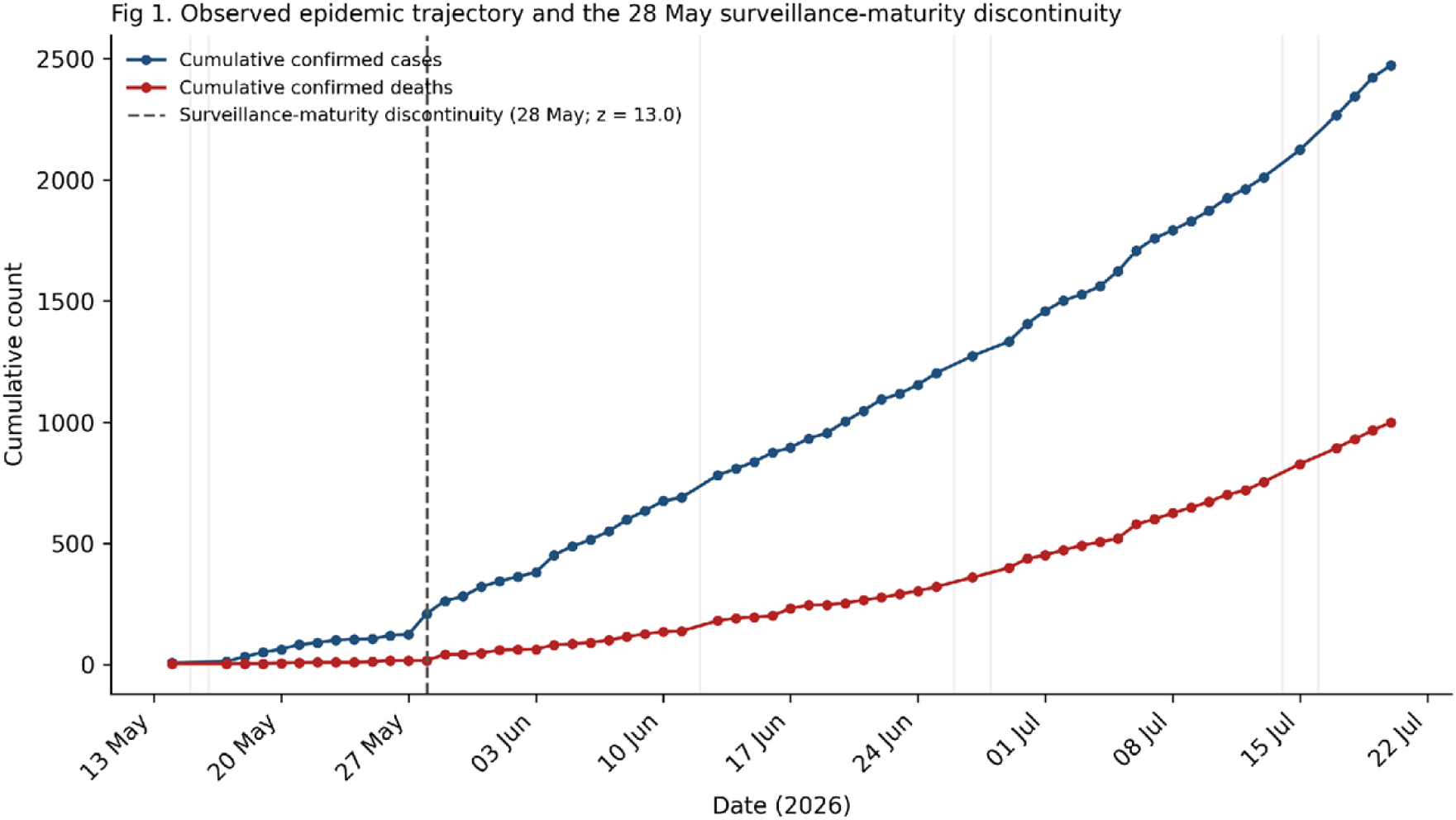
Observed epidemic trajectory, rolling forecast origins, and the 28 May surveillance-maturity discontinuity. Daily cumulative confirmed cases and deaths, missing-report dates (shaded), and the changepoint-detected discontinuity date, annotated with its robust z-score.

**Fig 2.**
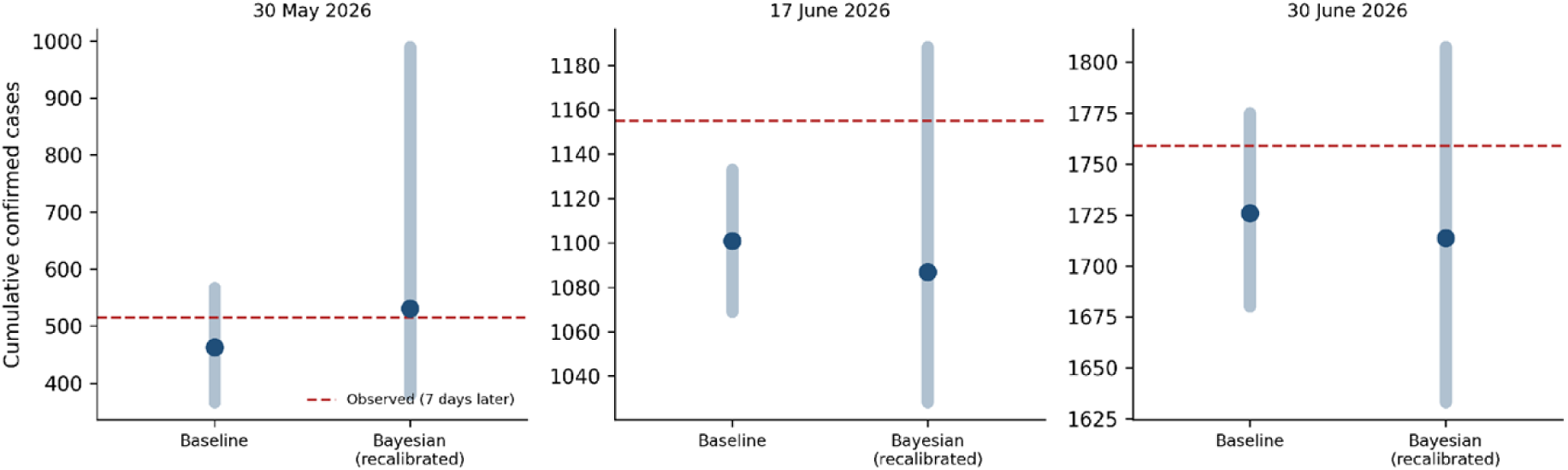
Prespecified rolling hindcasts. Three panels showing the information available on 30 May, 17 June, and 30 June 2026: baseline and recalibrated Bayesian forecasts (median, 80% interval) at the 7-day horizon, cumulative confirmed cases, with the subsequently observed total.

**Fig 3.**
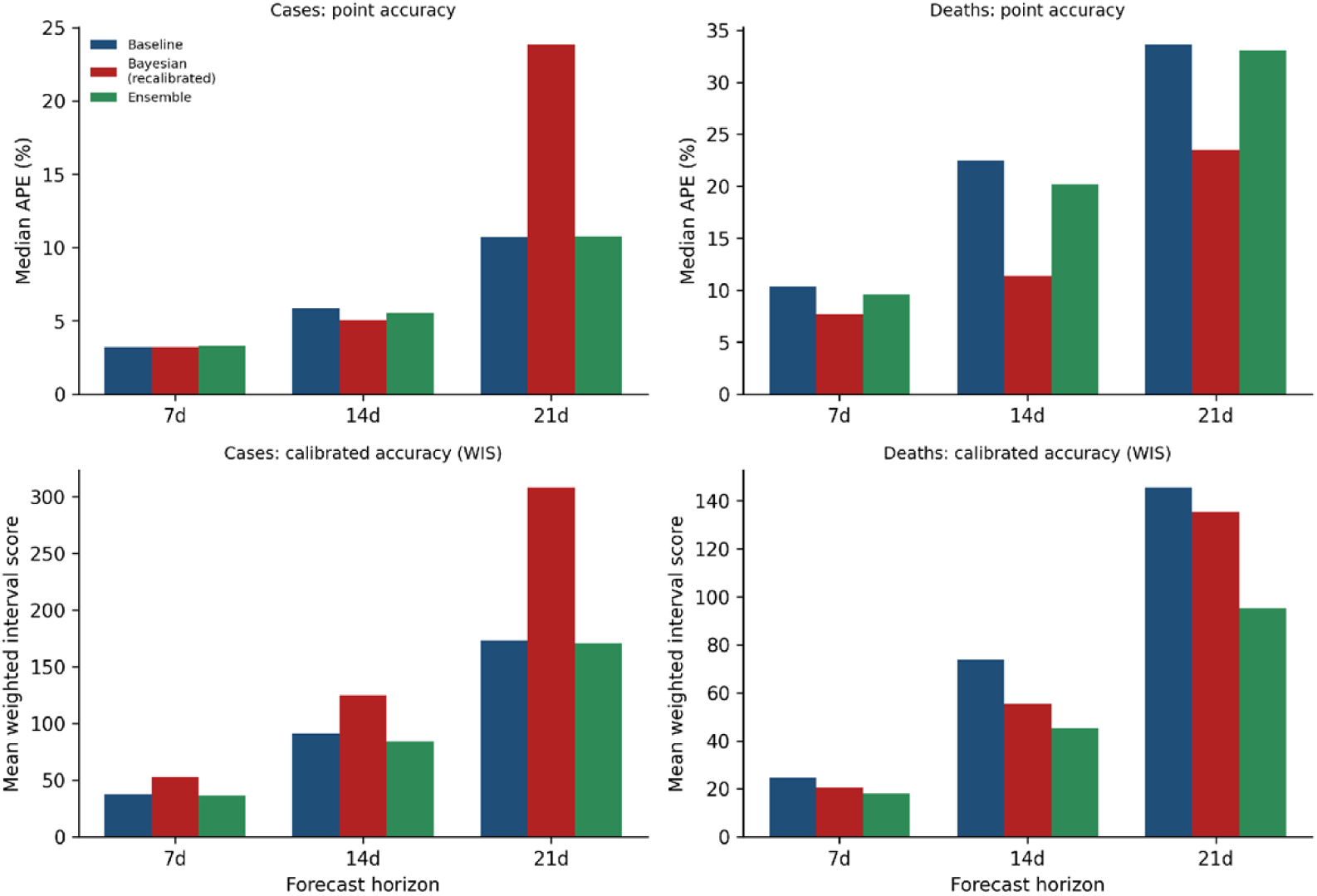
Forecast performance by model and horizon. Median absolute percentage error (point accuracy) and mean weighted interval score (calibrated accuracy) for confirmed cases and deaths at 7-, 14-, and 21-day horizons, data through 20 July 2026, for the baseline, recalibrated Bayesian model, and data-driven ensemble.

**Fig 4.**
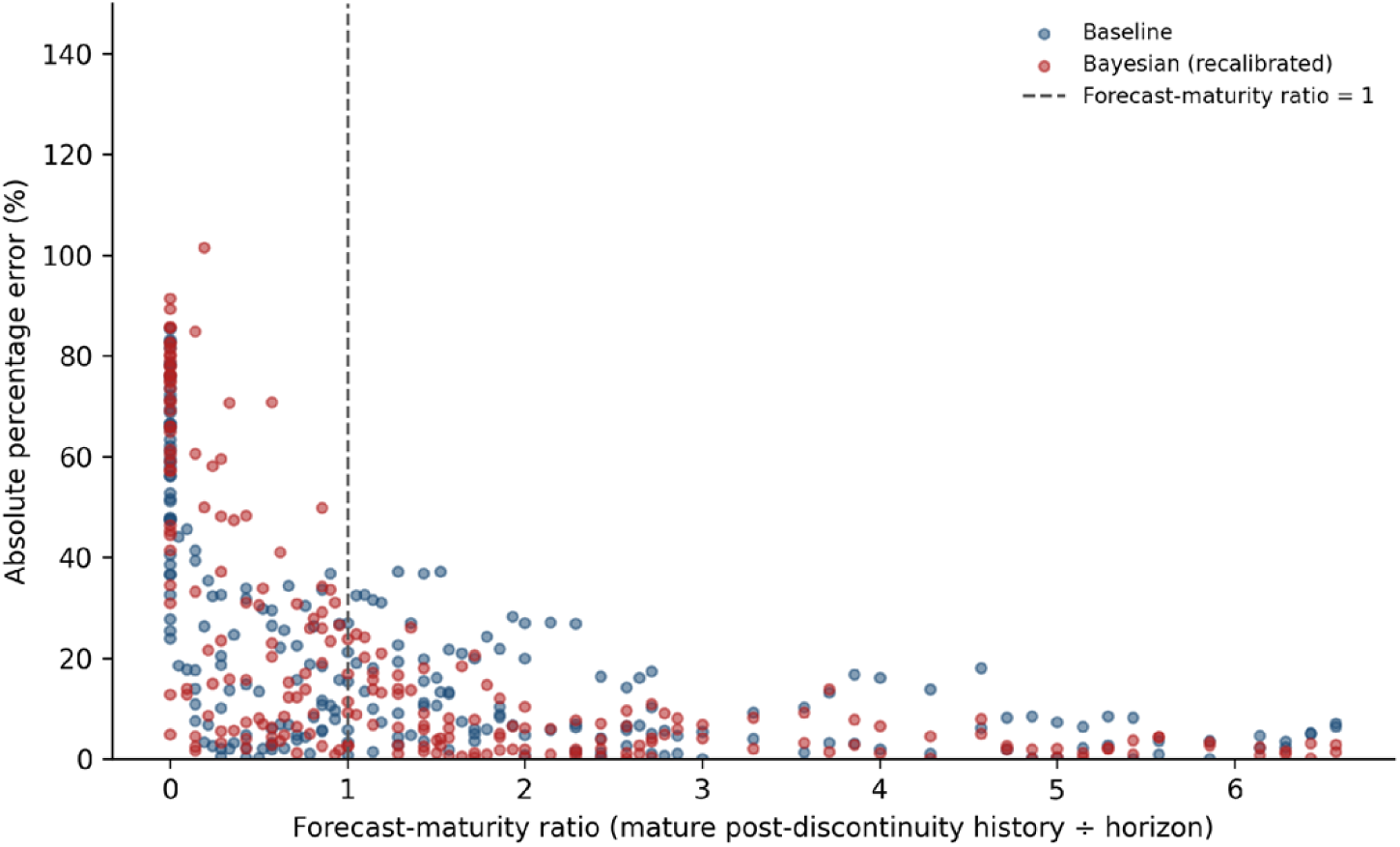
Forecast-maturity relationship, pooled across the baseline and recalibrated Bayesian models, 2026 BDBV dataset. [Note: the independent 2018–2020 North Kivu/Ituri external-validation panel from the original submission is not reproduced in this figure; the underlying ledger for that dataset was not part of this revision’s working files. It should be restored from the original archived pipeline before final submission.]

**Fig 5.**
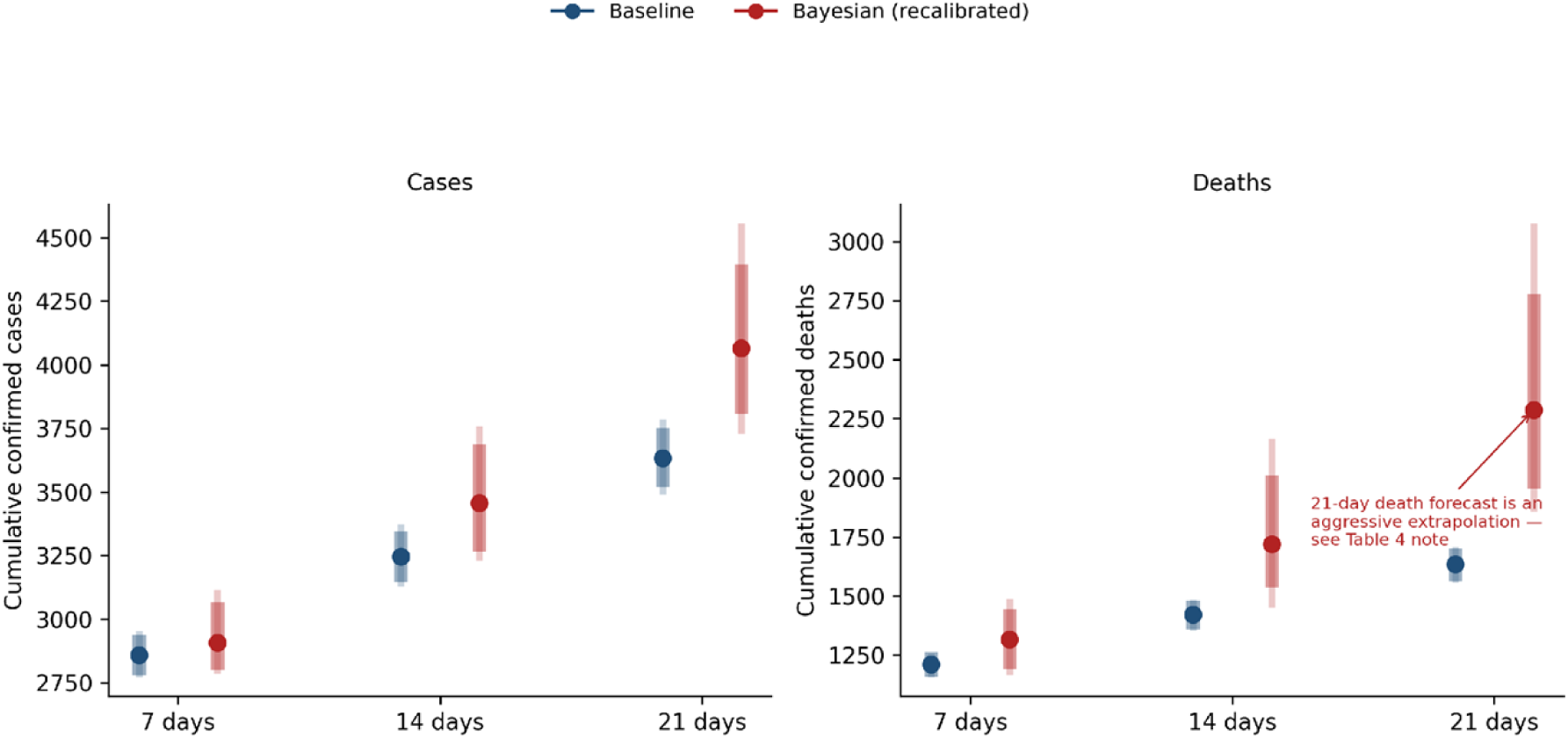
Prospective forecasts from the 20 July 2026 origin (median, 80% and 95% predictive intervals), baseline and recalibrated Bayesian model, cases and deaths, 7/14/21-day horizons. The 21-day death forecast is flagged as an aggressive extrapolation (Table 4). [Submission note: for final PLOS NTDs submission, each figure above must be uploaded as a separate standalone TIFF or EPS file (300–600 dpi, <10 MB), not embedded in the manuscript text; only the captions remain inline at the point of citation. The embedded figures above are provided in this version for internal review only.]

## Acknowledgments

We thank the INRB surveillance and situation-report teams whose routine reporting made this analysis possible.

## Data availability

All underlying surveillance data are publicly available without restriction from the INRB-UMIE BDBV2026-Data GitHub repository (https://github.com/INRB-UMIE/BDBV2026-Data, directory data/insp_sitrep/), which we accessed and archived on 20 July 2026 (repository commit hash and archive checksum retained by the corresponding author). The complete rolling-origin forecast ledgers, the empirical-recalibration and ensemble-weight-selection code, and the archived MCMC posteriors underlying every number reported in this paper will be deposited in a Zenodo repository at the time of acceptance and assigned a permanent DOI; a link will be added to this statement at that stage. No data are available only upon request.

## Author contributions

Conceptualization: JGLV, CNM, WJ. Data curation: JGLV. Formal analysis: JGLV. Methodology: JGLV, CNM. Software: JGLV. Validation: JGLV, CNM. Writing – original draft: JGLV. Writing – review & editing: all authors. Supervision: WJ.

## Generative AI use disclosure

During the preparation of this work the corresponding author used Claude (Anthropic) to assist with data validation, statistical pipeline implementation and rerunning, and drafting and revision of the manuscript text. After using this tool, the author reviewed and edited the content as needed and takes full responsibility for the content of the published article.

